# Elevated circulating CD16+ monocytes and TLR4+ monocytes in older adults with multiple cardiometabolic disease risk factors: A cross-sectional analysis

**DOI:** 10.1101/2021.04.13.21255420

**Authors:** Melissa M. Markofski, Michael G. Flynn

## Abstract

We endeavored to examine relationships between circulating monocyte phenotype and cardio-metabolic disease risk, in healthy, older adults. We performed a secondary data analysis on men and women, 55-75 yr, who were assigned to groups based on cardio-metabolic risk factors other than age. Subject in the low risk group (n=16, 12 females) had fewer than three risk factors. Subjects in the elevated risk group (n=29, 19 females) had three or more risk factors. Along with baseline screening for fitness and body composition, resting blood samples were assessed for markers of inflammation including: monocyte phenotype (inflammatory monocytes), monocyte cell-surface TLR4 expression, and serum C-reactive protein. The low risk group had a smaller (19.3% difference; p<0.0001) waist circumference and lower body fat weight (36.3%; p<0.0001), but higher 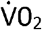max (45.5%; p=0.0019). There were no mean differences (p>0.05) between the low and elevated risk groups for BMI, serum cholesterol, fasting glucose, or leg press 1RM. The low risk group had lower CRP (114.7%, p=0.0002), higher CD14^+^CD16^-^ (classical) monocytes (6.7%; p=0.0231) and fewer CD14^+^CD16^+^ (inflammatory) monocytes (46.2%; p=0.0243) than the elevated risk group. The low risk group also had a lower percentage of CD14^+^CD16^+^ monocytes that were positive for TLR4 (14.0%; p=0.0328). Older men and women with fewer cardio-metabolic risk factors had lower serum and cellular markers of inflammation and higher aerobic capacity.

## 1. INTRODUCTION

Biomarkers of inflammation are linked to cardiometabolic diseases, for example type 2 diabetes and cardiovascular disease, such that people with cardiometabolic diseases have higher pro-inflammatory biomarkers than healthy individuals[1-5]. The hyper-inflammation may be both a product of the cardiometabolic diseases and a critical component of the development pathways for cardiometabolic diseases. For example, inflammation plays a role in the development of both insulin resistance and plaque development[6-8]. The link between inflammation and cardiometabolic disease is further supported by the observation that many of the medications to treat these diseases, such as metformin[9, 10], glitazones[11], and statins[12] have potent anti-inflammatory actions. Therefore, it is difficult to determine the proportion of these medication’s therapeutic actions that can be attributed to the reduction in inflammation or to other mechanisms.

There appear to be multiple pathways for the development of atherosclerosis. Once linked strongly to hyperlipidemia, pathways to atherogenesis now include the immune system in its pathology. In the lipid-centric model, lipids deposited in the arterial walls lead to upregulation of endothelium adhesion molecules[13]. These adhesion molecules promote monocyte attachment and transmigration, and the monocytes morph into pro-inflammatory foam cells that promote the vessel progression from fatty streak to atherosclerosis[6]. In contrast, the inflammation-initiated model features adhesion molecule-laden monocytes being recruited to the surface of the endothelium—in the absence of accumulated LDL. In this pathway, monocyte recruitment is believed to be facilitated by pro-inflammatory cytokines and pathogen-associated molecular patterns (PAMPs) or damage-associated molecular patterns (DAMPs), which initiate pro-inflammatory signaling[14-16].

It is clear that the lipid-centric model has dominated the attention of medical and pharmaceutical science, with effective medications available to lower LDL. However, since cholesterol lowering by itself does not preclude cardiac events, it is clear that the immune system’s role in these processes needs additional attention as it may also provide an as yet, undetermined therapeutic target[17, 18].

Central to both the lipid-centric and inflammation-centric models of atherosclerosis development is the involvement of monocytes, with mononuclear phagocytes playing a role in every step of the process. For example, significant murine inflammatory monocyte (Ly-6C^hi^) expansion was reported after high fat feeding, resulting in hypercholesterolemia, and these Ly-6C^hi^ cells were selectively accumulated in atherosclerotic lesions[19]. Following statin treatment, the Ly-6C^hi^ population contracted[19]. Inflammatory monocytes also have a role in cardiovascular disease in humans. Specifically, circulating inflammatory monocyte (CD14++CD16+) count at study enrollment accurately predicted cardiovascular events during the follow-up period[20].

Monocytes are a phagocytotic cell of the innate immune system with a broad array of cell-surface receptors, including adhesion molecules (VCAM), CD36 (oxLDL phagocytosis), and toll-like receptors, one of which, TLR4, confers LPS recognition on the monocyte[21]. However, there is evidence that high levels of TLR4 expression may be detrimental to health. For example, high expression of TLR4 is linked to poor health outcomes in a variety of conditions, including hepatitis C[22], atrial fibrillation[23], inflammatory bowel disease[24], and cancer.[25, 26] Specific to cancer, higher TLR4 expression on cancer cells is linked to reduced overall patient survival[26] and TLR4 influencing cancer cell survival after chemotherapy[25]. Further, it appears clear that TLR4 is implicated in the immune-centric model of atherosclerosis development[16]. Collectively, this supports that TLR4 may provide a useful target for new disease therapies.

The purpose of this study was to examine the relationship between cardiometabolic disease risk factors, monocyte phenotype, and monocyte TLR4 expression in healthy older adults. We hypothesized that older adults with three or more commonly measured cardiometabolic disease risk factors would have higher percentage of circulating monocytes and a greater proportion of monocytes positive for TLR4.

## 2. MATERIALS AND METHODS

### 2.1 Participants

As previously described[27], community dwelling men and women between the ages of 55-75 yrs were recruited for this study (females, n=29; males, n=16). Participants were free from chronic diseases (diabetes, osteoporosis, active cancer, cardiovascular disease with the exception of controlled blood pressure) known to impact study variables and from contraindications to exercise. Additionally, participants were not taking medication known to affect the immune system and/or the inflammatory response (β-blockers, statins, bisphosphonates, prescription anti-inflammatories, etc.). Current or recent users of tobacco products were excluded from participation. Finally, participants with high (>40 kg•m^2^) or low (<18 kg•m^2^) BMI were excluded. Data collection was approved by the Institutional Review Board at Purdue University.

### 2.2 Group assignment

Participants were assigned to a low risk for cardiovascular disease group (low risk; n=16, females n=12) or elevated risk for cardiovascular disease group (elevated risk; n=29, females n=19) based on a modified list of positive atherosclerotic cardiovascular disease risk factors and defining criteria[28]. Elevated risks were: age (women ≥55 yrs; men ≥45 yrs), physical inactivity (<3 d/wk of moderate to vigorous activity for the previous six months), overweight (BMI ≥25.0 kg•m^-2^), hyperlipidemia (total cholesterol ≥5.172 mmol/L), elevated fasting plasma glucose (≥5.5 mmol/L), waist circumference ≥88.0 cm for females or ≥100.0 cm for males, or C-reactive protein (CRP) ≥2.0 mg/L. As only adults over the elevated risk age were included in the study and this risk factor cannot be modified, all participants had at least one risk factor. Therefore, our threshold for “elevated” risk was age plus two modifiable risk factors (i.e., for the purpose of this study three risk factors or more was considered elevated risk). In addition, because of the independent effects of tobacco use on disease risk, participants were only included if they did not use tobacco products.

### 2.3 Screening

Prior to enrollment, participants were screened over the phone or via email for initial eligibility. If the participant met the initial eligibility criteria, they were scheduled for study visit 1. At this visit, the participant gave written informed consent and completed a self-reported medical history and physical activity history. Height and weight were measured to calculate BMI, and blood pressure measured after 15 min of seated rest. If the participant met all eligibility requirements, follow-up visits for fitness testing and body composition were scheduled.

### 2.4 Fitness testing and estimation of body composition

Briefly, all participants completed a two-stage treadmill test to estimate 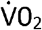max and 1 repetition maximum (1RM) testing on a pneumatic resistance leg press machine (Keiser, Fresno, CA, USA) to measure skeletal muscle strength. Prior to the leg press test, participants who reported they were not currently strength training completed familiarization sessions on three non-consecutive days. Participants arrived in the morning after an overnight fast for relative body composition estimation with a BodPod (Cosmed, Concord, CA, USA). The relative body composition measurement was used to estimate fat weight and fat-free weight. Waist circumference was measured using a Gulick tape measure.

### 2.5 Blood sample collection and analyses

As previously described[27], participants arrived to our laboratory after an overnight fast and refraining from exercise for the previous 72 hr. After a 15-minute seated rest, blood was collected from a vein in the antecubital space. Sodium-heparinized blood was used for same-day flow cytometry analyses, and serum was collected and stored at -80°C for batch analyses of circulating biomarkers. Three-color flow cytometry (CD14, CD16, TLR4) was used to determine monocyte phenotype. Glucose and cholesterol was measured using commercially available endpoint assay kits (Infinity, Thermo-Fisher) and CRP was analyzed using a commercially available ELISA kit (ALPCO, Salem, NH, USA).

### 2.6 Statistical design and analyses

This is a secondary data analyses of a subset of participants in a completed study[27]. All participants with complete data for the eight risk factors were included in these analyses. Sample size was calculated on group differences in CD14+CD16+ from a previous exercise training study in older adults[29], to achieve 80% power with alpha = 0.05. Prior to analyses, data were evaluated for normality equal variance, and transformed prior to ANOVA analyses. All data are presented non-transformed as mean ± standard error. Analysis of variance models were fit by risk group. All statistical analyses were conducted with statistical software (SAS v9.4, Cary, NC, USA).

## 3. RESULTS

Participants in the low cardiovascular disease risk factors group had lower waist circumference (p<0.0001), CRP (p=0.0002), fat-free weight (p<0.0001), and higher (p=0.0019) 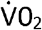max than participants in the elevated cardiovascular disease risk factors group (Table 1). There were no differences (p>0.05) between groups for age, BMI, cholesterol, glucose, fat free weight, or leg press 1RM (Table 1).

**Table 1:** Participant characteristics for subjects in the low risk (n=16) and elevated risk (n=29) group. * Significant = p < 0.05.

|  | **Low**<br>n=16/12<br>(75%<br>female) | **Elevated**<br>n=29/19<br>(66%female) | **p value** |
| --- | --- | --- | --- |
| Age (yrs) | 58.3±1.0 | 58.3±1.2 | 0.8254 |
| BMI ($\text{kg} \cdot \text{m}^{-2}$) | 26.0±0.7 | 31.5±0.7 | 0.3272 |
| Waist circumference (cm) | 82.4±1.9 | 99.0±2.2 | <0.0001* |
| Cholesterol (mmol/L) | 4.71±0.2 | 5.1±0.2 | 0.3272 |
| Glucose (mmol/L) | 5.4±0.1 | 5.7±0.1 | 0.3825 |
| CRP (mg/L) | 1.3±0.2 | 4.8±.8 | 0.0002* |
| Fat-free weight (kg) | 49.2±2.9 | 54.0±2.4 | 0.2079 |
| Fat weight (kg) | 25.5±2.2 | 36.8±1.6 | <0.0001* |
| $\dot{V}O_{2\text{max}}$ (est.) | 41.1±6.4 | 25.9±1.4 | 0.0019* |
| Leg Press 1RM (N) | 172.0±15.2 | 188.3±11.1 | 0.3423 |

Participants in the elevated cardiovascular disease risk group had a lower (p=0.0231) percentage of circulating classical (CD14+CD16-) monocytes, and higher (p=0.0243) percentage of circulating inflammatory (CD14+CD16+) monocytes (Figure 1). Participants in the elevated cardiovascular disease group also had a higher (p=0.0328) percentage of classical (CD14+CD16-) monocytes positive for TLR4. There was no group difference (p>0.05) in percentage of inflammatory (CD14+CD16+) monocytes positive for TLR4 (Figure 2).

**Figure 1.**
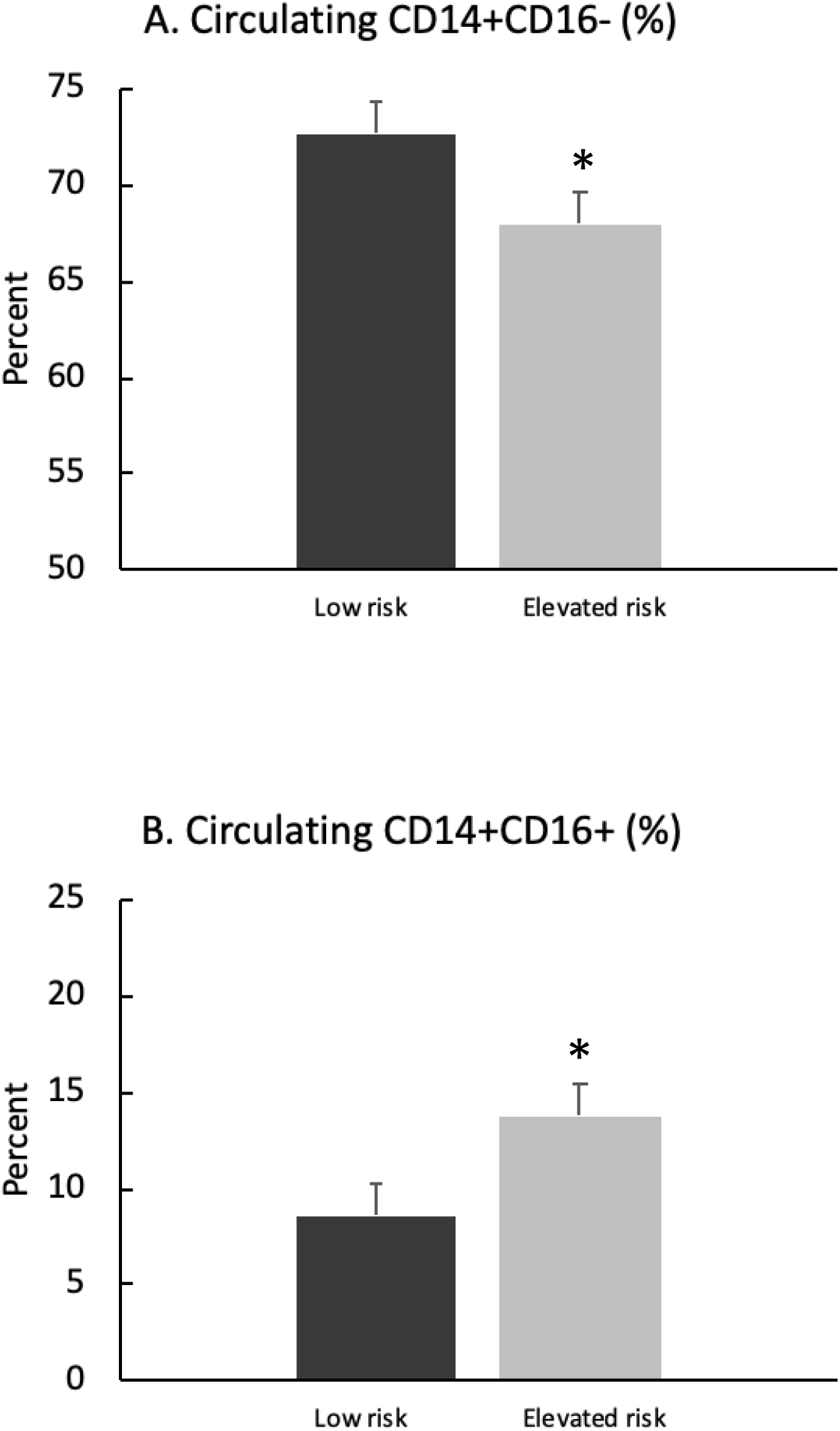
Circulating monocyte phenotypes. Mean ±S.E. for percent circulating CD14+CD16-monocytes (1A) and CD14+CD16+ monocytes (1B) for subjects in low risk group (black bar) and elevated risk group (gray bar). *p<0.05

**Figure 2.**
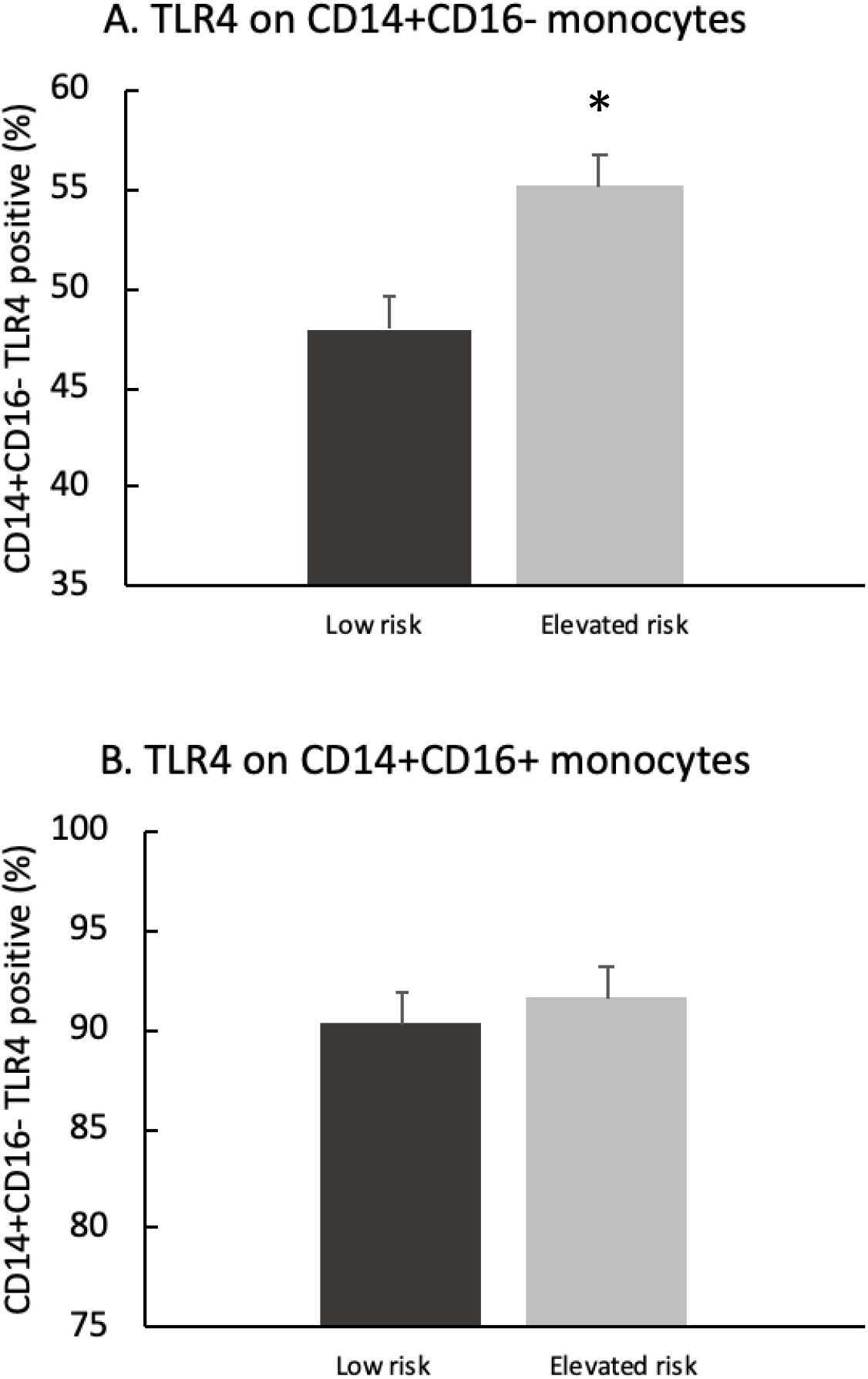
Percent of monocytes positive for TLR4 on CD14+CD16-(2A) monocytes and CD14+CD16+ monocytes (2B) for subjects in low risk group (black bar) and elevated risk group (gray bar). *p<0.05

## 4. DISCUSSION

In this secondary analysis we have assessed the importance of established cardiometabolic risk factors for determining immune biomarkers associated with cardiometabolic risk. Older, overweight non-tobacco users with an elevated risk for cardiovascular disease had higher percentages of circulating monocytes associated with increased inflammation and disease risk. Specifically, the elevated risk group had higher percentages of circulating inflammatory (CD14+CD16+) monocytes and of circulating classical monocytes (CD14+CD16-) positive for TLR4. There was no difference in the percentage of inflammatory monocytes positive for TLR4. Our data add to the growing body of literature supporting the importance of inflammatory markers in chronic disease or, in this case, the risk factors for chronic disease in an otherwise healthy tobacco-free population of older adults. As an example, the focus on lipid parameters in cardiovascular disease is important, but has gaps for identifying risk or targeting treatment as a large proportion of patients with heart disease do not “fit” the lipidcentric model[18]. We, and others, have demonstrated that the circulating percentage of inflammatory monocytes and other inflammatory biomarkers respond favorably to exercise[27, 30-34], providing a simple, low-cost, and apparently effective influence on an important therapeutic target.

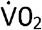max was higher and CRP lower in the low risk group compared to the group with three or more cardiometabolic risk factors (elevated risk). Higher estimated 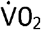max, used as a proxy for physical activity levels, agrees with our earlier research that physically active adults had lower markers of inflammation than physically inactive subjects[31]. We have also previously reported that improving 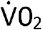max via training has similar blunting effects on TLR4 and inflammatory monocyte expression[30]. The present study did not divide participants by self-reported physical activity level because self-reported physical activity level was one of the included risk factors. It is interesting to note that although 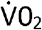max was higher in the low risk group, self-reported physical activity level was distributed between the two groups.

Participants with three or more risk factors had a higher proportion of circulating inflammatory monocytes (CD14+CD16+), but low circulating classical monocytes (CD14+CD16-). Inflammatory monocytes are linked to plaque formation[35], intimal wall thickening in pre-hypertension[36], and are correlated with a wide range of cardiovascular risk factors and predictive of myocardial infarction, non-hemorrhagic stroke, or cardiovascular death[20]. Specifically, Rogacev et al.[20] reported a difference in event-free survival between patients in the highest and lowest quartiles of inflammatory monocytes. Further, the impact of monocyte count varied depending on model and number of covariates. It has also long been known that inflammatory monocytes and macrophage infiltration contribute to the formation of vascular plaques; however, the infiltration is often independent of cholesterol levels as was the case for diabetic patients in the Burke et al. study[37]. We hypothesized that cardiometabolic risk factors would be predictive of these cellular and biomarkers and, similar to Rogacev et al.[20], found that patients with more risk factors have higher CRP, circulating percentage of inflammatory monocytes, and cell-surface TLR4.

Toll-like receptor expression on a variety of cell types has been linked to chronic disease[38-40]. TLR4 expression is found to promote inflammation associated with these diseases[38, 40]. In the present study, we found higher TLR4 expression on CD14+16-monocytes in subjects from an elevated risk group compared to a lower risk group of older subjects. However, there were no differences between these groups for TLR expression on CD14+16+ monocytes. A simple explanation for the latter is that the activated, pro-inflammatory monocytes are highly expressive of TLR4, in general, with both groups above 90% of monocytes expressing TLR4. Thus, a ceiling effect may have influenced our ability to detect differences between the groups. Nevertheless, the elevated expression of TLR4 in vascular plaques[14] suggests an important relationship that was not revealed in these activated CD14+16+ monocytes.

One strength of this study is that all participants were relatively healthy at the time of the study, and did not report taking medications known to influence the immune system and inflammatory biomarkers. Studying participants who are in good health but have differences in immune biomarkers linked to chronic diseases may aid in explaining why some people who appear to be in good health develop chronic diseases, whereas others do not. One limitation of the study is that it only included participants of a specific, older age range and not all adults. A future study could repeat the analyses in a wider age range.

## 5. CONCLUSION

In summary, older men and women with fewer cardiometabolic risk factors had lower serum and cellular markers of inflammation and higher aerobic capacity. Specifically, the participants with fewer cardiometabolic risk factors had a lower percentage of circulating inflammatory CD14+CD16+ monocytes, and a lower number of circulating classical CD14+CD16-monocytes positive for cell surface TLR4. These results strengthen the growing support for the role of inflammation and TLR4 in chronic diseases.

## Supporting information

STROBE checklist

## Data Availability

Data available upon request to corresponding author

## Acknowledgements

MGF: This research was supported (in whole or in part) by HCA and/or an HCA affiliated entity. The views expressed in this publication represent those of the author(s) and do not necessarily represent the official views of HCA or any of its affiliated entities.

